# WEPP: Phylogenetic Placement Achieves Near-Haplotype Resolution in Wastewater-Based Epidemiology

**DOI:** 10.1101/2025.06.09.25329287

**Authors:** Pranav Gangwar, Pratik Katte, Manu Bhat, Yatish Turakhia

## Abstract

Wastewater carries the full spectrum of pathogens and their variants infecting a population, making it a powerful resource for public health surveillance. During the SARS-CoV-2 pandemic, wastewater-based epidemiology (WBE) proved its value by providing a cost-effective and unbiased means to detect emerging variants days to weeks ahead of clinical reporting, driving its widespread global adoption. However, wastewater remains a relatively untapped resource for genomic epidemiology because most computational tools for WBE are limited to lineage-level resolution, focusing only on estimating the abundance of different lineages from wastewater sequencing reads. Here, we present WEPP, a pathogen-agnostic pipeline that significantly enhances the resolution and capabilities of WBE analysis. WEPP uses phylogenetic placement of wastewater sequencing reads onto comprehensive phylogenies—specifically, mutation-annotated trees (MATs) that include all globally available clinical sequences and their inferred ancestral nodes—to identify a subset of haplotypes most likely present in the sample. In addition, WEPP reports the abundance of each haplotype and its corresponding lineage, provides parsimonious mappings of individual reads to haplotypes, and flags ‘unaccounted alleles’—those observed in the sample but unexplained by selected haplotypes–that may signal the presence of novel circulating variants. Using over 100 simulated, synthetic, and real-world SARS-CoV-2 wastewater samples, we demonstrate that WEPP not only surpasses existing lineage abundance tools in terms of accuracy but also achieves near-haplotype-level resolution, typically selecting haplotypes that are within an average distance of one single-nucleotide mutation from the true haplotype. This level of resolution overcomes key limitations of current WBE approaches and enables new applications that were previously confined to clinical sequencing, such as tracking intra-lineage haplotype clusters, identifying the geographical origins of newly introduced clusters, and detecting emerging variants. We further demonstrate WEPP’s generalizability by applying it to wastewater samples of two additional pathogens. WEPP also includes an interactive visualization dashboard that supports unprecedented high-resolution analysis, allowing users to visualize detected haplotypes and haplotype clusters within the context of the global phylogenetic tree, investigate haplotype and lineage abundances, examine alignments of reads parsimoniously mapped to selected haplotypes, and inspect unaccounted (cryptic) alleles. With these capabilities, WEPP has the potential to transform wastewater-based epidemiology into a more powerful tool for investigating and managing infectious disease outbreaks.

**Code availability:** The WEPP source code is freely available under the MIT license at https://github.com/TurakhiaLab/WEPP, with comprehensive documentation to support new users available at https://turakhia.ucsd.edu/WEPP.

## Introduction

Wastewater carries a comprehensive mixture of pathogens shed by infected individuals in a catchment area, making wastewater-based epidemiology (WBE) a powerful tool for monitoring community-level pathogen dynamics that is also non-invasive, cost-effective, timely, and unbiased^1^. During the COVID-19 pandemic, WBE demonstrated successful applications in detecting emerging SARS-CoV-2 lineages up to two weeks before clinical reporting^2–6^, enabling timely public health interventions. This success has driven the widespread adoption of WBE across more than 55 countries^7^, including many low- and middle-income countries (LMICs), with deployments ranging from university dormitories^2,8,9^ to hospitals^10–12^ and airports^13–15^. It has also catalyzed its expansion to several other pathogens, including influenza, respiratory syncytial virus (RSV), rhinovirus, monkeypox (Mpox), Zika, and Human papillomavirus (HPV)^16–19^.

Despite immense success, substantial opportunities remain untapped in WBE, as, to date, most WBE tools have focused predominantly on estimating the proportions of various lineages^2,20–27^. As a result, several critical epidemiological applications that necessitate *haplotype-level resolution* remain reliant on clinical sequencing. These applications include: (i) tracking intra-lineage haplotype clusters to study local transmission dynamics^28,29^, (ii) identifying introductions of haplotype clusters into a region^30^, and (iii) detecting emerging variants that are not yet designated with lineage labels^31,32^. Since clinical sequencing rates have been in sharp decline since the official end of the COVID-19 pandemic^33^, it is imperative that we empower WBE with more capabilities, particularly by improving its resolution, as we transition to a broader post-pandemic surveillance era, as well as expand WBE to broad pathogens.

While there have been prior efforts^20^ to reconstruct haplotypes directly from sequencing reads, these have only been applied to mixed samples containing haplotypes from two very distinct clusters (separated by 200 to 400 mutations^20^), and they struggle to reconstruct haplotypes within clusters exhibiting low genetic diversity (separated by fewer than 100 mutations). As a result, these approaches are not well-suited for WBE samples^34^, which often feature complex mixtures of dozens of haplotypes with low genetic diversity. Their performance is further exacerbated when applied to short-read sequencing data, which remains the predominant sequencing technology in WBE applications^2,22,24–26,34^. Pipes et al.^35^ proposed an alternative approach to overcome these challenges–rather than inferring haplotypes from scratch, their method leverages an expectation-maximization (EM) algorithm to select the most likely subset of haplotypes from a database of previously sequenced clinical samples, thus providing improved resolution over lineage abundance estimation tools. However, even this method is suited only for samples containing a relatively small number (around 10) of unique haplotypes^35^, and the computationally intensive nature of the EM algorithm necessitates aggressive pre-filtering strategies that impact its accuracy (see Results). Moreover, this method does not allow the discovery of novel or ‘cryptic’ variants^36–41^ that have not yet been observed in clinical sequencing data.

To address these shortcomings, we introduce WEPP (Wastewater-based Epidemiology using Phylogenetic Placements)–a pathogen-agnostic pipeline that significantly improves the accuracy and resolution of wastewater-based epidemiology (Figure 1A). Specifically, WEPP places sequencing reads onto mutation-annotated trees (MATs)—which are comprehensive, up-to-date phylogenies maintained by the UShER toolkit^42,43^—to identify likely haplotypes (Figure 1A) from global clinical and inferred ancestral sequences, corresponding to the tip and internal nodes of the MAT, respectively. WEPP then uses a deconvolution algorithm (modified from Freyja^2^) to estimate the relative abundances of the selected haplotypes and reports ‘unaccounted alleles’—alleles present in wastewater but unexplained by the selected haplotypes. These unaccounted alleles closely resemble the ‘cryptic’ mutations described in prior studies^36,37,40,41^, which are often indicative of a potentially novel circulating variant. However, unaccounted alleles may also arise from systematic sequencing or bioinformatic errors, or suboptimal haplotype selection (Results). To further enhance the analysis, WEPP includes an interactive visualization dashboard that enables users to explore detected haplotypes within the context of a comprehensive phylogenetic tree consisting of all globally available clinical sequences (Figure 1B). WEPP’s dashboard also displays the relative abundances of different haplotypes and their corresponding lineages, alignments of reads parsimoniously assigned to the selected haplotypes, and unaccounted alleles (Figure 1B). Such fine-grained analysis of wastewater samples is currently not possible with available tools and dashboards, which can empower new applications in WBE.

**Figure 1:**
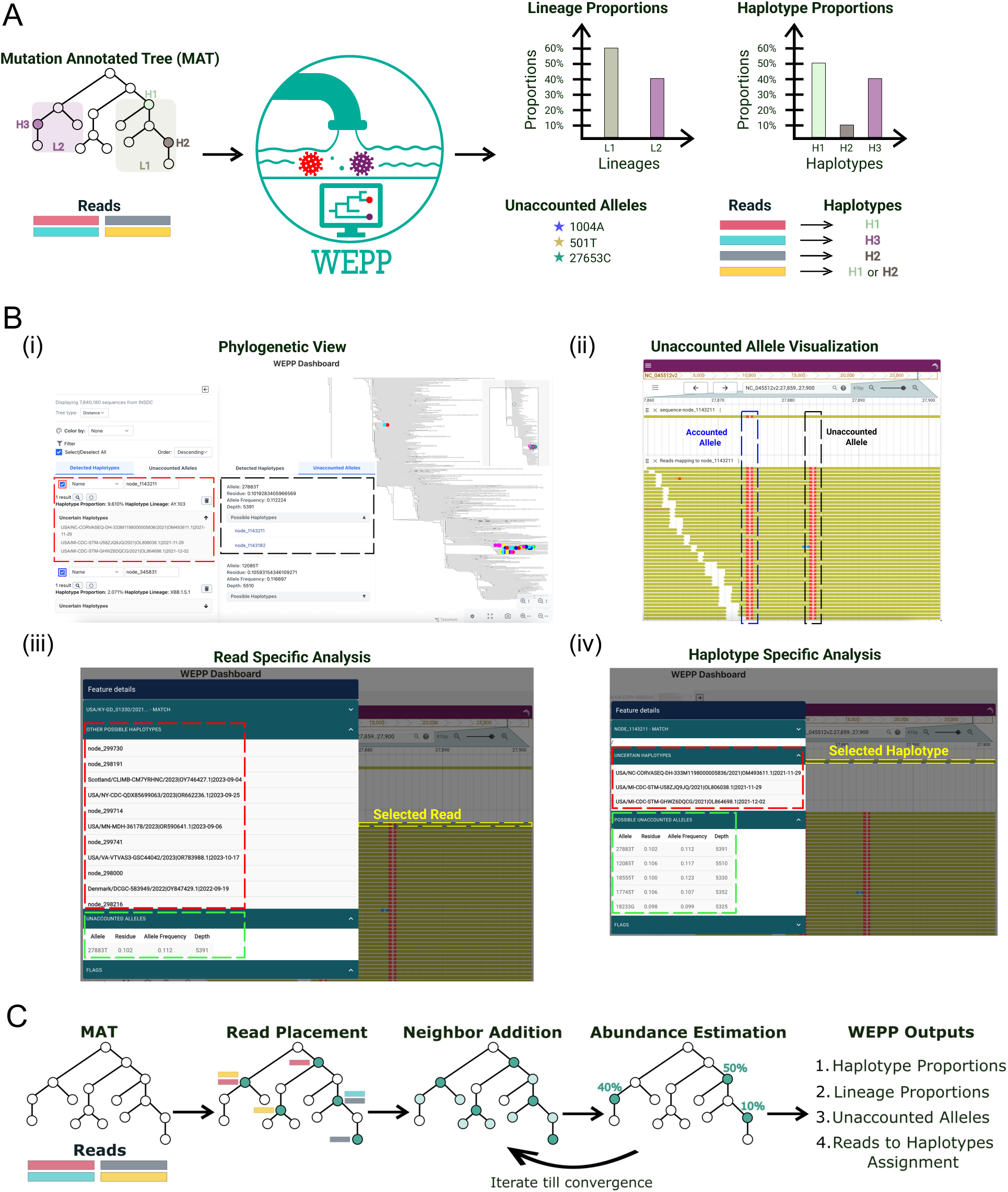
Overview of the WEPP pipeline. (A) WEPP input and output. (B) Features of the interactive WEPP Dashboard: (i) Phylogenetic View of the WEPP inferred haplotypes along with their estimated proportions, associated lineages, and any uncertain haplotypes; (ii) Read analysis panel highlighting accounted and unaccounted alleles contained in reads mapped to a selected haplotype: the first highlighted site shows an alternate allele present in both the reads and the haplotype, while the second shows an alternate allele present in the reads but absent from the haplotype; (iii) Read information panel displaying all possible haplotypes and unaccounted alleles for a selected read; (iv) Haplotype information panel listing the possible unaccounted alleles associated with the selected haplotype. (C) Key stages of WEPP’s phylogenetic algorithm for haplotype detection and abundance estimation.

Through our extensive evaluation using diverse benchmarks across multiple pathogens, we demonstrate WEPP’s excellent performance across a wide range of 54 simulated, 13 synthetic (laboratory-engineered), and 77 real-world wastewater samples. Notably, for the lineage abundance estimation task on simulated and synthetic SARS-CoV-2 wastewater samples, where ground truth is available, WEPP reduced root-mean-square error (RMSE) by 5-fold and 2.4-fold, respectively, compared to state-of-the-art methods. Moreover, in each of these datasets, WEPP sensitively and precisely identified the likely haplotypes from a search space of millions in the SARS-CoV-2 MAT, with the inferred haplotypes being, on average, at a distance of less than one mutation from the actual haplotype. WEPP also accurately identified all intra-lineage haplotype clusters present in the samples. We further demonstrate that WEPP can be reliably used to detect cluster introductions in different regions before their appearance in clinical sequencing, and flag mutations of the novel variants, which are still clinically undetected. When applied to real SARS-CoV-2 wastewater samples from Point Loma (San Diego), WEPP exhibited a higher correlation with clinical sequencing data than a state-of-the-art tool and detected multiple independent Omicron introductions into the city, identifying several haplotypes in the wastewater samples that were later clinically confirmed. Additionally, the unaccounted alleles reported by WEPP from the Point Loma samples, as well as from real wastewater samples described in Suarez et al.^36^, showed strong overlap with high-confidence cryptic mutations. WEPP also performed robustly on hospital wastewater samples from Columbia University^10^, which were sequenced using long-read technology from Oxford Nanopore Technologies (ONT), showing strong correlation with the hospital’s clinical sequencing data. Using RSV-A and Mpox samples, including real-world RSV-A wastewater samples from Geneva and Zurich, we also demonstrate WEPP’s generalizability across diverse pathogens, achieving high accuracy and consistent identification of lineages missed by clinical sequencing.

Overall, WEPP provides an automated, accurate, scalable, and flexible framework that enables near-haplotype-level resolution, thereby opening new avenues for genomic surveillance of pathogens and significantly broadening the scope and utility of wastewater-based epidemiology in the post-pandemic era.

## Results

### WEPP Overview

WEPP is a phylogeny-based pipeline for WBE that works complementarily with clinical sequencing efforts. WEPP takes as input raw wastewater sequencing reads and a mutation-annotated tree (MAT) containing clinical sequences of the pathogen being analyzed, and it outputs a set of haplotypes (MAT nodes) with their estimated abundances that best explain the wastewater sample (Figure 1A). WEPP also reports lineage abundances, unaccounted alleles, and parsimonious read-to-haplotype mappings (Figure 1A)—all of which can be visualized through an interactive, user-friendly dashboard (Figure 1B). By enabling near-haplotype-level resolution within the context of a comprehensive global phylogeny built from clinical sequencing data, WEPP extends the capabilities of wastewater-based epidemiology (WBE) to a broader range of applications. These include: (i) detecting intra-lineage clusters circulating within the local catchment area, (ii) inferring introductions of new transmission clusters across regions, (iii) identifying unaccounted alleles that may indicate emerging variants, and (iv) performing detailed, read-level analysis of the wastewater sample.

Figure 1C provides a high-level overview of the WEPP’s core algorithm, with a detailed description available in the Methods section. Briefly, WEPP starts by performing parsimonious placement of raw sequencing reads on the mutation-annotated tree (MAT), using an optimized version of the UShER algorithm tailored for the phylogenetic placement of sequencing reads rather than whole-genome sequences (Methods). Based on the resulting placement distribution, WEPP scores and selects a subset of haplotypes, along with their nearest neighbors (around 5000 total haplotypes, Methods), to form a pool of candidate haplotypes. This pool is passed to a deconvolution algorithm, which is based on Freyja^2^, to estimate the relative abundance of each node. Since Freyja’s formulation does not include a regularization term, to mitigate overfitting, WEPP only retains nodes above a user-defined abundance threshold (default: 0.5%) and iteratively refines this set. In each iteration, a new candidate pool is formed from the current haplotypes above this frequency threshold and their corresponding neighbors (default: 2 mutation radius; maximum 500 neighbors per haplotype), followed by deconvolution. This process continues until it reaches convergence or a maximum iteration count (default: 10). WEPP uses an outlier detection algorithm on the residue of the deconvolution algorithm, suggesting a wide gap between observed and estimated allele frequencies, to flag a list of unaccounted alleles (Methods). WEPP incorporates several optimizations to achieve pandemic-scale analysis, allowing it to place in minutes to hours millions of wastewater sequencing reads on SARS-CoV-2 MATs that consist of tens of millions of nodes (Methods).

WEPP’s interactive dashboard can be launched as a local web server through WEPP’s Snakemake pipeline (Methods) and is designed to enhance interpretability and support in-depth downstream analysis. As shown in Figure 1B(i), the dashboard presents a detailed phylogenetic view, similar to Taxonium^44^, of all clinically sequenced haplotypes, with the nodes corresponding to the ones identified in the wastewater sample by WEPP highlighted in colored circles. The dashboard also displays the estimated abundances of each node, their associated lineages, and any uncertainty in node selection caused by uncovered sites in the sample. Users can also perform a detailed read-level analysis by selecting a haplotype (MAT node) to view its characteristic mutations alongside those observed in the mapped reads (Figure 1B(ii)). Additional information about individual reads or haplotypes can be accessed by clicking on their corresponding objects, as shown in Figures 1B(iii) and 1B(iv), respectively.

### WEPP significantly improves lineage abundance estimation accuracy and achieves near-haplotype resolution on simulated and synthetic wastewater data

We first sought to determine whether WEPP can accurately estimate lineage abundances and resolve haplotypes circulating in the wastewater. This assessment used a combination of simulated and synthetic wastewater datasets with known ground truth (Methods). The synthetic wastewater samples were obtained from the Indiana Department of Health (IDOH) Laboratory and the data shared by Ferdous et al.^45^, who sequenced their samples on Illumina and Oxford Nanopore Technologies (ONT) platforms, respectively.

The simulation analysis was performed on 10 samples using SWAMPy^46^, a realistic wastewater simulation tool, in which the lineage abundances were made to resemble those reported by CA-SEARCH surveillance efforts (https://searchcovid.info) between December 2022 and December 2023, and the haplotypes within those lineages were chosen randomly (Methods). To quantify the accuracy of lineage abundances and compare the results of WEPP to Freyja^2^ and Pipes et al.^35^, we used the Root Mean Square Error (RMSE) between expected and estimated abundances. To quantify the ability of WEPP to resolve haplotypes, we introduce two new metrics: i) Weighted Haplotype Distance and ii) Weighted Peak Distance (Methods). Briefly, Weighted Haplotype Distance quantifies how closely wastewater haplotypes match WEPP’s inferred haplotypes (referred to as “peaks” to avoid confusion with wastewater haplotypes), where a lower value implies higher *sensitivity*. At the same time, Weighted Peak Distance assesses how well-estimated peaks align with true haplotypes in the sample, where a lower value implies higher *precision*. Since Freyja does not report actual haplotypes, we used root sequences corresponding to Freyja’s reported lineages as their representative peaks for comparison.

We observed that WEPP significantly outperformed both Freyja and Pipes et al. on the most widely performed lineage abundance estimation task, achieving nearly 5-fold and 7.9-fold lower RMSE on average compared to Freyja and Pipes et al., respectively (Figure 2A). A more detailed sample-wise analysis (Supplementary Table 1) reveals that because Freyja only optimizes over phylogenetically-derived mutational barcodes for different lineages, it often struggles with neighboring lineages (e.g., EG.5 and EG.5.1 in the October 2023 sample, Supplementary Table 1), where a haplotype from a particular lineage circulating in the wastewater sample shares some mutations with its neighboring lineage. Similarly, WEPP significantly outperforms the technique proposed by Pipes et al., which is known to struggle with samples containing more than 10 haplotypes^35^, whereas our simulated samples carried up to 100 haplotypes. Both Freyja and Pipes et al. report far more lineage than what is present in the wastewater sample (Supplementary Table 1). Overall, WEPP benefits not only from searching over all haplotypes in the MAT, but also by including the inferred sequences at the internal nodes of the MAT, allowing it to more precisely identify the haplotypes circulating in the wastewater and their phylogenetic contexts on the full phylogeny, thus leading to more reliable estimates. Moreover, WEPP exhibits strong sensitivity and precision in identifying haplotypes circulating in wastewater, achieving a 3.1-fold and 6.4-fold reduction in Weighted Haplotype Distance compared to Freyja and Pipes et al., respectively. Similarly, it attains a 6.9-fold and 7.5-fold lower Weighted Peak Distance relative to Freyja and Pipes et al. (Figure 2A and Supplementary Table 1). In fact, the WEPP distance values are close to zero, indicating that most WEPP-inferred haplotypes perfectly match the true haplotypes present in the sample.

**Figure 2:**
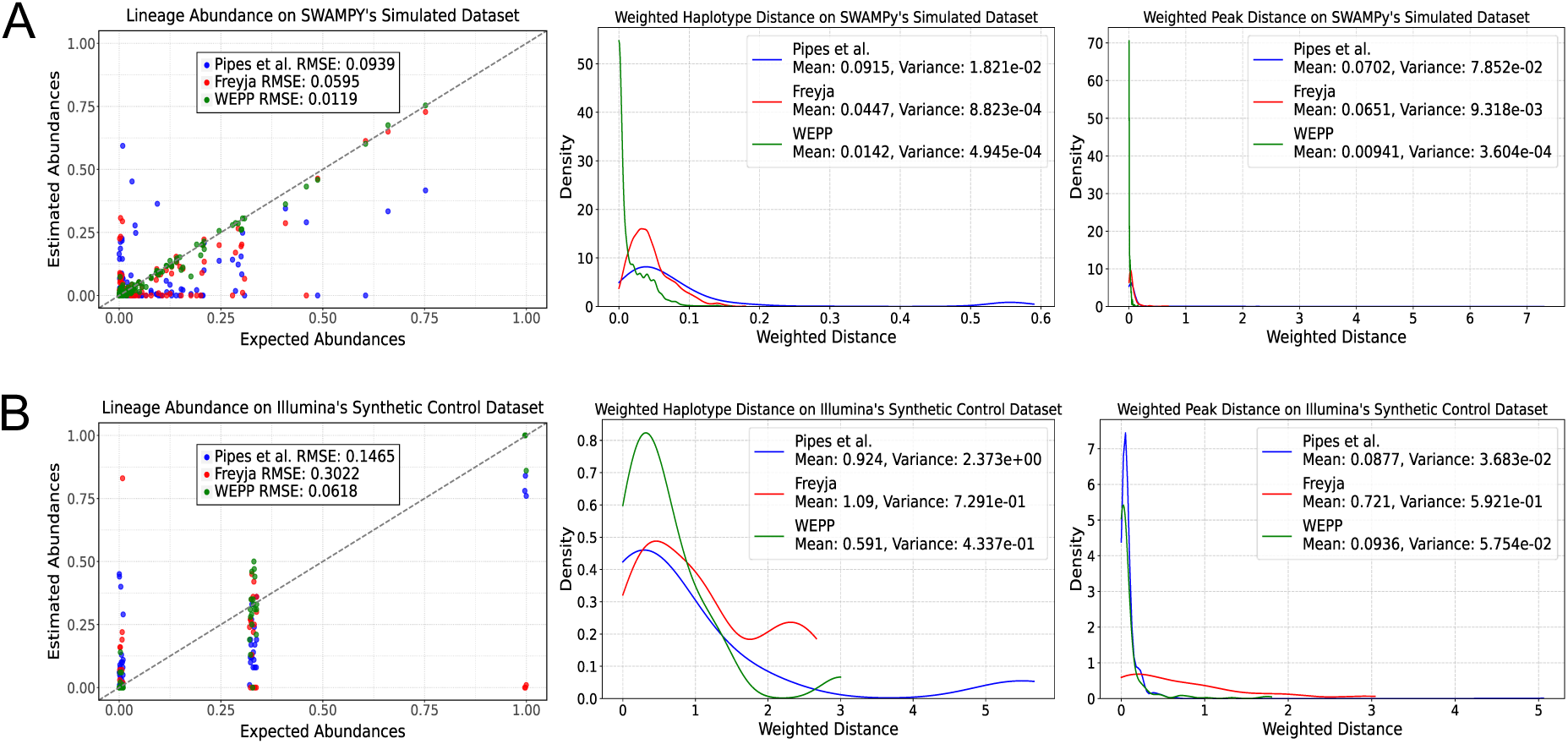
Evaluation of lineage abundance accuracy and haplotype-level resolution in WEPP, Freyja, and Pipes et al. across simulated and synthetic control mixtures. (A) Comparison of Lineage Abundance estimates, Weighted Haplotype Distance, and Weighted Peak Distance on SWAMPy simulated datasets. (B) Comparison of lineage abundance estimates, Weighted Haplotype Distance, and Weighted Peak Distance on the Illumina sequenced synthetic control datasets.

We further evaluated all three tools over ten Illumina-sequenced synthetic control mixtures produced and sequenced by the Indiana Department of Health (IDOH) Laboratory, where synthetic viral RNA from known haplotypes was added to the wastewater to mimic real samples (Methods). Consistent with earlier findings, WEPP achieved substantial improvements over Freyja, with 4.9-fold lower lineage abundance RMSE, 1.8-fold lower Weighted Haplotype Distance, and 7.7-fold lower Weighted Peak Distance. The technique of Pipes et al. likely benefited from the simplicity of the synthetic samples, which included only 1–3 haplotypes, enabling their method to achieve a 1.1-fold lower Weighted Peak Distance than WEPP, indicating marginally better precision. However, WEPP performed significantly better on remaining metrics, achieving 2.4-fold and 1.6-fold reductions on RMSE and Weighted Haplotype Distance, respectively, compared to Pipes et al. (Figure 2B and Supplementary Table 1).

We observed similar trends when evaluating WBE tools on three synthetic wastewater samples from Ferdous et al.^45^, sequenced using high-error long reads (ONT). WEPP achieved a 3.2-fold reduction in RMSE compared to Freyja (Supplementary Figure 1A and Supplementary Table 1). Despite the elevated error rates in ONT reads, WEPP’s inferred haplotypes remained close to the ground truth, highlighting its robustness and generalizability.

Overall, WEPP’s strong performance on both simulated and synthetic wastewater samples across two distinct sequencing technologies underscores its ability to enhance the accuracy and resolution of wastewater surveillance.

### WEPP can accurately identify intra-lineage circulating clusters

The increased resolution of WEPP’s analysis at the haplotype level has the potential to unlock powerful downstream applications for epidemiologists using wastewater data. During much of the COVID-19 pandemic, local transmission at any given time point was often driven by just a few dominant viral lineages^2,47–49^. These broad lineage-level trends could be effectively tracked using existing wastewater-based epidemiology (WBE) tools, such as Freyja. However, within many of these lineages lies substantial genetic diversity, e.g., two haplotypes belonging to the BA.1 lineage could differ by as many as 50 substitutions (Supplementary Figure 2), which can mask important epidemiological dynamics. For example, distinct co-circulating clusters within a lineage may represent multiple independent introductions of the virus into a region or the emergence of localized outbreak events, and public health officials have typically needed to turn to clinical sequencing data to detect these patterns^50–54^.

We evaluated whether WEPP can accurately detect intra-lineage haplotype clusters by simulating wastewater samples containing clinical sequences detected in San Diego during the months of January and September 2023. These months were chosen to capture diverse variants while ensuring a sufficient number of available sequences. Using “CA-SEARCH,” we identified the different haplotypes clinically sequenced in San Diego, simulated wastewater samples containing all haplotypes from a month using SWAMPy, and compared them to the WEPP’s detected haplotypes (Figure 3A-B).

**Figure 3:**
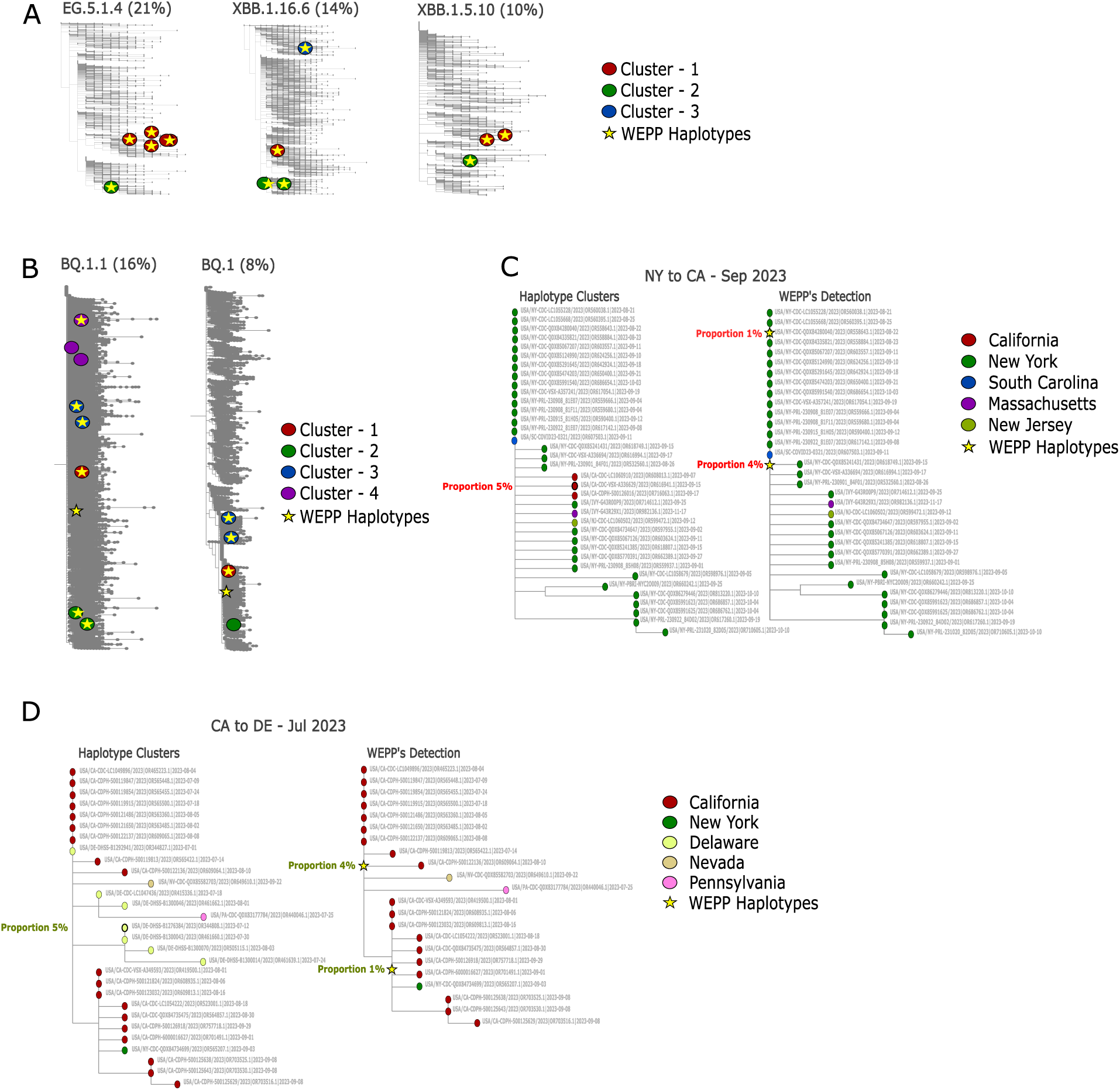
Demonstrating WEPP’s haplotype detection capabilities across various scenarios, with visualizations made with the help of WEPP’s dashboard. (A) WEPP’s detection of intra-lineage haplotype clusters circulating in San Diego in September 2023. (B) WEPP’s detection of intra-lineage haplotype clusters circulating in San Diego in January 2023. (C) Left: ClusterTracker indicates a likely variant introduction from New York to California. Right: WEPP’s detected haplotypes in the New York-origin cluster using a MAT excluding California haplotypes. (D) Left: ClusterTracker indicates a likely variant introduction from California to Delaware. Right: WEPP’s detected haplotypes in the California-origin cluster using a MAT excluding Delaware haplotypes.

$$$Figure 3A shows that in September 2023, the EG.5.1.4, XBB.1.5.10, and XBB.1.16.6 lineages collectively made up 45% of detected sequences in San Diego, while in January 2023, BQ.1.1 and BQ.1 accounted for 24% of all sequences (Figure 3B). Many of these lineages contained haplotype clusters with distinct genetic makeups (e.g., BQ.1.1 clusters were separated by up to 28 substitutions as shown in Supplementary Figure 2), which existing WBE tools, like Freyja, would not be able to discern. While Freyja generally estimated the lineage proportions accurately (Supplementary Table 2), WEPP also identified all haplotypes in the September 2023 dataset (Figure 3A) and detected most circulating clusters in January 2023 (Figure 3B).

By resolving this intra-lineage variation directly from wastewater samples, WEPP could provide a complementary lens for real-time surveillance and finer-scale tracking of pathogen spread, especially in settings with limited clinical sequencing capacity.

### WEPP can enable early detection of new cluster introductions from wastewater sequencing

Previous studies have demonstrated that wastewater surveillance can detect emerging viral lineages up to two weeks before clinical sequencing^2^. While this capability has underscored the utility of wastewater surveillance as an early warning system, current WBE tools cannot investigate the introductions and origins of emerging variant clusters in different geographical locations. Specifically, while this feature exists for clinical sequencing data through tools like ClusterTracker^30^– a tool widely used by public health officials in the US. We are not aware of any counterpart that is based on wastewater data.

Similar to ClusterTracker, WEPP’s increased resolution has the potential to flag the introduction of new variant clusters in a geographical region and track their likely source of origin, but using wastewater sequencing data. We used simulated data to evaluate this capability since such a level of ground truth is not available for any real-world data. Specifically, we simulated cluster emergence of two transmission events discovered by ClusterTracker: (1) New York to California transmission of an EG.5.1.9 variant cluster in September 2023 (Figure 3C), and (2) California to Delaware transmission of an HU.1.1 variant cluster in July 2023 (Figure 3D). Detailed methodology for simulating cluster emergence in wastewater is provided in Methods.

On both datasets, WEPP demonstrated promising results. Since the actual haplotypes that were introduced in the new region (California and Delaware, respectively) were removed from the input MAT, WEPP instead detected two closely related haplotypes at 4% and 1% abundance in both cases— belonging to (1) the New York originating cluster in the first experiment (Figure 3C), and (2) the California originating cluster in the second (Figure 3D). Moreover, in both cases, the haplotypes surrounding the WEPP’s detected haplotypes belonged to their probable sources—New York and California, respectively—strengthening confidence in the inferred transmission pathways. These results underscore WEPP’s potential to detect emerging haplotype clusters in a region and identify their possible sources of origin, possibly even before they are detected through clinical sequencing. This opens a new application for wastewater sequencing, thereby expanding its role in public health surveillance.

### WEPP can identify mutations present in novel variants

Timely detection of new variants is essential for guiding public health responses and can help save lives^55,56^. Delays in detecting emerging variants from clinical sequencing are well documented^57,58^; moreover, clinical sequencing rates declined sharply following the end of the Public Health Emergency declaration in May 2023^33^, which has impacted the ability to track new variants effectively. In contrast, due to its cost-effectiveness, wastewater surveillance programs remain active in many parts of the world and are therefore playing an increasingly important role in monitoring variants. While previous wastewater studies have identified early cryptic SARS-CoV-2 variants that were unexplained by clinical sequencing^36,37,38^, they have typically involved manual efforts and were limited in scope (e.g., focusing on rare, co-occurring mutations), which, as our results later indicate, could be prone to accuracy and sensitivity issues.

As described earlier, WEPP extends Freyja’s deconvolution framework to identify alleles whose observed frequencies from wastewater deviate significantly from expectations based on the inferred haplotypes and their abundances. These deviations, referred to as unaccounted alleles, may signal cryptic mutations—potential early indicators of emerging variants or novel lineages. To assess the sensitivity and precision of this approach, we systematically varied the abundance of a novel haplotype and its genetic divergence from known sequences in the phylogenetic tree (indicated by ‘Node Removal Radius’, Table 1). As evident in the results presented in Table 1, increasing the genetic divergence of the novel haplotype from known sequences makes it more challenging to detect its closest match in the MAT. This is reflected by the distance between the novel haplotype and the nearest haplotype estimated by WEPP, which exceeds the minimum value defined by the ‘Node Removal Radius’ at higher divergence. Nevertheless, WEPP consistently recovered all mutations associated with highly divergent haplotypes, even at low proportions. Moreover, nearly every unaccounted allele flagged by WEPP corresponded to ‘Distance-reducing Alleles’ (Methods)—alleles that, if assigned to the correct inferred haplotype, would reduce dissimilarity to the true sequences in the sample. As the abundance of the novel haplotype increased, the number of unaccounted alleles declined, consistent with WEPP’s adaptive outlier detection strategy that becomes more stringent in the presence of stronger signals (Methods). These findings highlight WEPP’s ability to accurately detect private mutations and identify the closest phylogenetic matches for novel haplotypes, even when present at only 5% abundance.

**Table 1:** Evaluation of WEPP’s Unaccounted Alleles and closest haplotype from novel haplotypes in the phylogenetic tree. WEPP’s sensitivity and precision in detecting novel haplotypes are assessed by (1) varying the novel haplotype’s proportion in the sample, and (2) adjusting the mutation distance between the novel haplotype and its closest sequence in the tree, defined by the ‘Node Removal Radius’. The columns represent: (i) the novel haplotype’s proportion, (ii) ‘Node Removal Radius’, (iii) the number of unaccounted alleles detected by WEPP, (iv) the number of mutations that reduce the distance between WEPP inferred haplotypes and the wastewater haplotypes, and (v) the mutation distance between the novel haplotype and the closest haplotype detected by WEPP.

| Averaged results over four datasets |  |  |  |  |
| --- | --- | --- | --- | --- |
| Novel Haplotype Proportion | Node Removal Radius | Distance between the novel haplotype and the closest WEPP's estimated haplotype | Unaccounted Alleles | Distance-reducing Alleles |
| 20% | 3 | 3 | 3 | 3 |
|  | 5 | 6.5 | 6.5 | 6.5 |
|  | 7 | 8 | 8.25 | 8 |
| 10% | 3 | 3 | 8 | 8 |
|  | 5 | 6.5 | 7.25 | 7.25 |
|  | 7 | 7.5 | 8 | 8 |
| 5% | 3 | 3 | 9 | 8.25 |
|  | 5 | 6.5 | 10.5 | 10 |
|  | 7 | 8 | 10.5 | 10 |

We further validated this capability by analyzing 22 SARS-CoV-2 wastewater samples from the Point Loma wastewater treatment plant (San Diego) between November 27, 2021, and February 7, 2022, and comparing them against 12,908 clinical sequences reported in San Diego during the peak surveillance period from November 2021 to March 2022 (Supplementary Table 6, Methods). Of the 330 unaccounted alleles flagged by WEPP, 35.8% (118 alleles) were completely absent from the clinical sequences, and an additional 43.0% (142 alleles) were extremely rare, present in fewer than 1% of clinical sequences. According to prior definitions^37,38^, these novel or clinically rare alleles would typically be categorized as ‘cryptic mutations’. In other words, the unaccounted alleles identified by WEPP have a high overlap (nearly 79%) with cryptic mutation definition, whose origins may stem from undetected low-prevalence variants or upstream processing artifacts. The remaining 21% unaccounted alleles, although present in clinical data, were flagged by WEPP because their observed frequencies in the wastewater sample deviated substantially from the expected frequencies based on haplotype selection, suggesting either the accumulation of previously observed mutations in circulating variants or systematic errors in sequencing or bioinformatics pipelines.

To evaluate WEPP’s potential for cryptic lineage detection, we analyzed a subset of the Suarez et al.^36^ datasets, which identified consensus sequences for several cryptic lineages. They identified “cryptic lineage-defining amino acid substitutions” (cryptic mutations) from prior studies and searched for sequencing reads containing multiple such co-occurring mutations, and applied additional custom filtering criteria. We analyzed the 104 cryptic mutations reported by Suarez et al. across 18 samples (Supplementary Table 9) and found that WEPP flagged 26% of them as unaccounted alleles. The remaining mutations were not flagged because 49% fell below WEPP’s read-depth threshold, 20.2% fell below the depth-weighted allele-frequency cutoff, and 4.8% were already captured within WEPP’s inferred haplotypes. In other words, mutations not flagged by WEPP do not carry a “strong” signal to be confidently classified as cryptic. These results underscore that because WEPP integrates global phylogenetic context alongside stringent allele frequency and sequencing depth thresholds, it ensures that only high-confidence cryptic mutations are reported as unaccounted alleles. While this approach may differ from earlier strategies, we believe it provides a principled and balanced framework for maximizing both sensitivity and specificity in cryptic mutation detection.

Together, these findings demonstrate that by reporting unaccounted alleles, WEPP offers critical insights that can help public health officials assess emerging or anomalous patterns in pathogen evolution and uncover novel and cryptic variants with high precision and sensitivity.

### WEPP closely aligns with local clinical sequences on real wastewater data

We compared WEPP’s lineage detection accuracy against Freyja using both short-read, low-error Illumina-sequenced wastewater samples from the Point Loma wastewater plant (San Diego) and long-read, high-error ONT-sequenced hospital wastewater surveillance data from Annavajhala et al.^10^, with contemporaneous clinical sequencing data serving as a benchmark. Clinical sample collection was simultaneously conducted at both locations, which typically precedes public database deposition by a median of 10 to 63 days^59^, but serves as a high-confidence “oracle” reference for our comparisons. Our first evaluation focused on 22 Illumina-sequenced wastewater samples collected between November 27, 2021, and February 7, 2022, from the Point Loma Wastewater Treatment Plant, during a period of highest clinical sequencing efforts in the city. Additionally, we analyzed 13 long-read ONT-sequenced wastewater samples collected in December 2022 from Columbia University’s hospital, described in Annavajhala et al., and compared them against 57 clinical sequences collected from the same hospital during that month. Lineage abundance estimates from wastewater were evaluated against clinical data to assess the sensitivity and concordance of both tools with circulating SARS-CoV-2 lineages.

A key distinction between the tools was Freyja’s detection of a substantially greater number of lineages across both datasets (Supplementary Tables 3 and 4). However, many of these lineages were inferred at low relative abundances and lacked corresponding support from clinical sequencing, even during the peak surveillance period in San Diego (Figure 4, Supplementary Table 4). As reflected in our controlled lineage abundance experiments (Figure 2), these lineages indicate a certain degree of overfitting in Freyja’s estimations. To quantify the concordance between wastewater-inferred and clinical lineage distributions, we performed Kullback-Leibler (KL) divergence analysis, comparing Point Loma wastewater estimates to those from “oracle” clinical sequencing (applying a one-week shift to account for earlier detection via wastewater). WEPP’s estimates consistently exhibited lower divergence with clinical data (Figure 4C), highlighting improved precision and sensitivity in detecting circulating variants.

**Figure 4:**
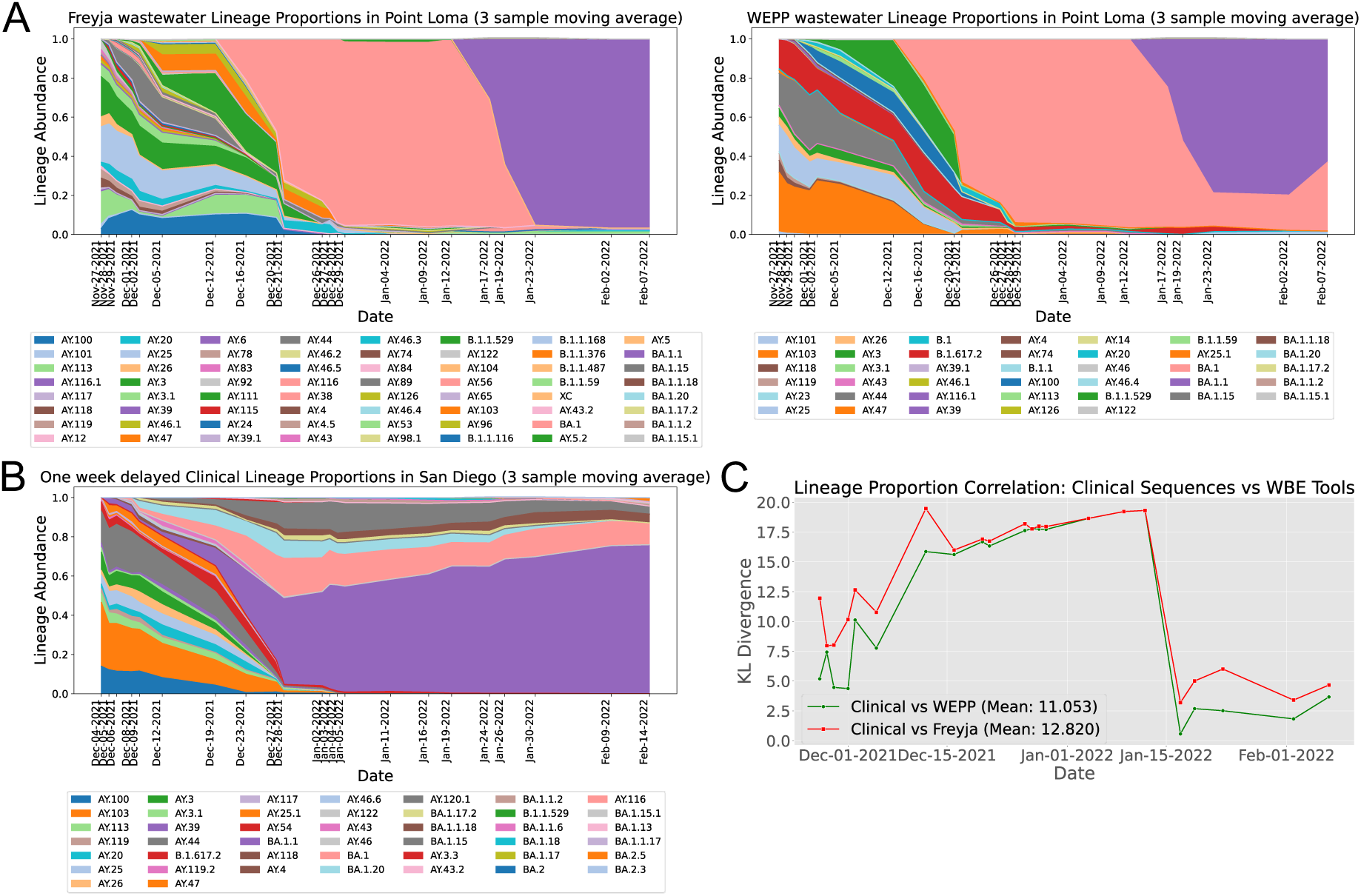
Comparison of wastewater-based lineage estimates with one-week delayed clinical sequences. (A) Lineage proportion estimates from Freyja (left) and WEPP (right) for Point Loma wastewater samples (Nov 27, 2021 – Feb 7, 2022). (B) Lineage proportions derived from one-week delayed “Oracle” clinical sequences in San Diego during the same period. (C) KL divergence between one-week delayed clinical sequence-derived lineage proportions and those estimated from WEPP and Freyja from wastewater.

Both WEPP and Freyja detected the Omicron variant (B.1.1.529) in wastewater as early as December 1, 2021, using the MAT from the same date that included some Omicron sequences. Each tool reported the variant at approximately 2% relative abundance (Supplementary Table 4). Notably, WEPP identified two distinct Omicron haplotypes at 0.9% abundance in that sample. One haplotype was later confirmed by 127 clinical sequences from San Diego beginning December 8, 2021, while the other was observed clinically only three times, starting January 8, 2022. In total, 32 haplotypes identified by WEPP in wastewater were also observed in the clinical data (Supplementary Table 5). More importantly, WEPP detected more than one distinct haplotype cluster within the B.1.1.529 (Omicron) lineage in wastewater samples as early as December 1, 2, and 5, 2021, revealing at least two independent Omicron introductions into San Diego (Supplementary Table 10 and Supplementary Figure 3). By identifying these separate introduction events at low proportions and at least a week before clinical confirmation, WEPP could have guided targeted interventions (e.g., focused testing, enhanced contact tracing) to curb the spread.

We further evaluated the impact of MAT updates on the detection accuracy of both tools by simulating a possible pandemic setting in which UShER MATs are updated slowly, every 15 days. With the MAT update for samples collected post-December 15, 2021, both WEPP and Freyja detected BA.1 at over 60% relative abundance, while prior samples had no estimation of BA.1 due to the absence of BA.1 haplotypes in the older MAT from December 1, 2021 (Figure 4A). Similarly, before January 15, 2022, both WEPP and Freyja detected only the BA.1 lineage (Figure 4A), as the earlier MATs (dated December 15, 2021, and January 1, 2022) did not contain any BA.1.1 haplotypes. However, following the MAT update for samples collected after January 15, 2022, WEPP successfully identified both BA.1 and BA.1.1 lineages in wastewater, whereas Freyja detected only BA.1.1. Retrospective analysis of clinical sequences from San Diego, which were unavailable at the time due to reporting delays^57^, confirms that each of BA.1 and BA.1.1 accounted for more than 15% of cases from mid-December onward, with BA.1.1 increasing in prevalence over time—a trend more closely mirrored by WEPP’s results (Figure 4B). This underscores WEPP’s enhanced sensitivity in resolving closely related lineages from real wastewater data, which would enable epidemiologists to more accurately track pathogens’ transmission and evolutionary dynamics.

To assess WEPP’s performance under noisy sequencing conditions, we also analyzed high-error, long-read ONT-sequenced wastewater samples from Columbia University’s hospital, described in Annavajhala et al., collected in December 2022. For both WEPP and Freyja, we averaged the estimated lineage abundances across all samples and computed the KL divergence against lineage abundances from clinical sequences collected during the same month. The KL divergence values were comparable for both tools (Supplementary Figures 1B and 1C, Supplementary Table 3), demonstrating that WEPP maintains robust performance under noisy sequencing conditions.

Collectively, these results highlight WEPP’s robustness across sequencing technologies, demonstrating high sensitivity and accuracy towards emerging variant detection, even at very low abundances, and identifying multiple local introductions at least a week ahead of clinical confirmation. This early signal could provide critical lead time for public health officials to respond and mitigate potential large-scale outbreaks. Its enhanced lineage detection sensitivity also uncovers low-frequency transmission events, while the unaccounted alleles provide early warning of cryptic transmissions that may drive future caseloads. These findings underscore that WEPP would enable more effective and timely interventions, and also demonstrate the necessity of frequent MAT updates for sustaining high accuracy in wastewater-based surveillance.

### WEPP generalizes across pathogens and detects lineages missed by clinical surveillance

Wastewater-based surveillance is increasingly being expanded to track a range of infectious diseases beyond SARS-CoV-2, including Influenza A, RSV, Mpox, Measles, and Hepatitis. However, unlike SARS-CoV-2, none of these pathogens are extensively sequenced, which places greater importance on WBE tools that can generalize well and fully leverage all the available sequence data.

To demonstrate WEPP’s pathogen-agnostic capabilities, we evaluated its lineage estimation capabilities on two other pathogens: Respiratory Syncytial Virus Subgroup A (RSV-A) and Mpox, using three simulated datasets for each pathogen (Figure 5A; see Methods). WEPP accurately detected the abundances of all the expected lineages in both cases, validating its applicability across diverse pathogens.

**Figure 5:**
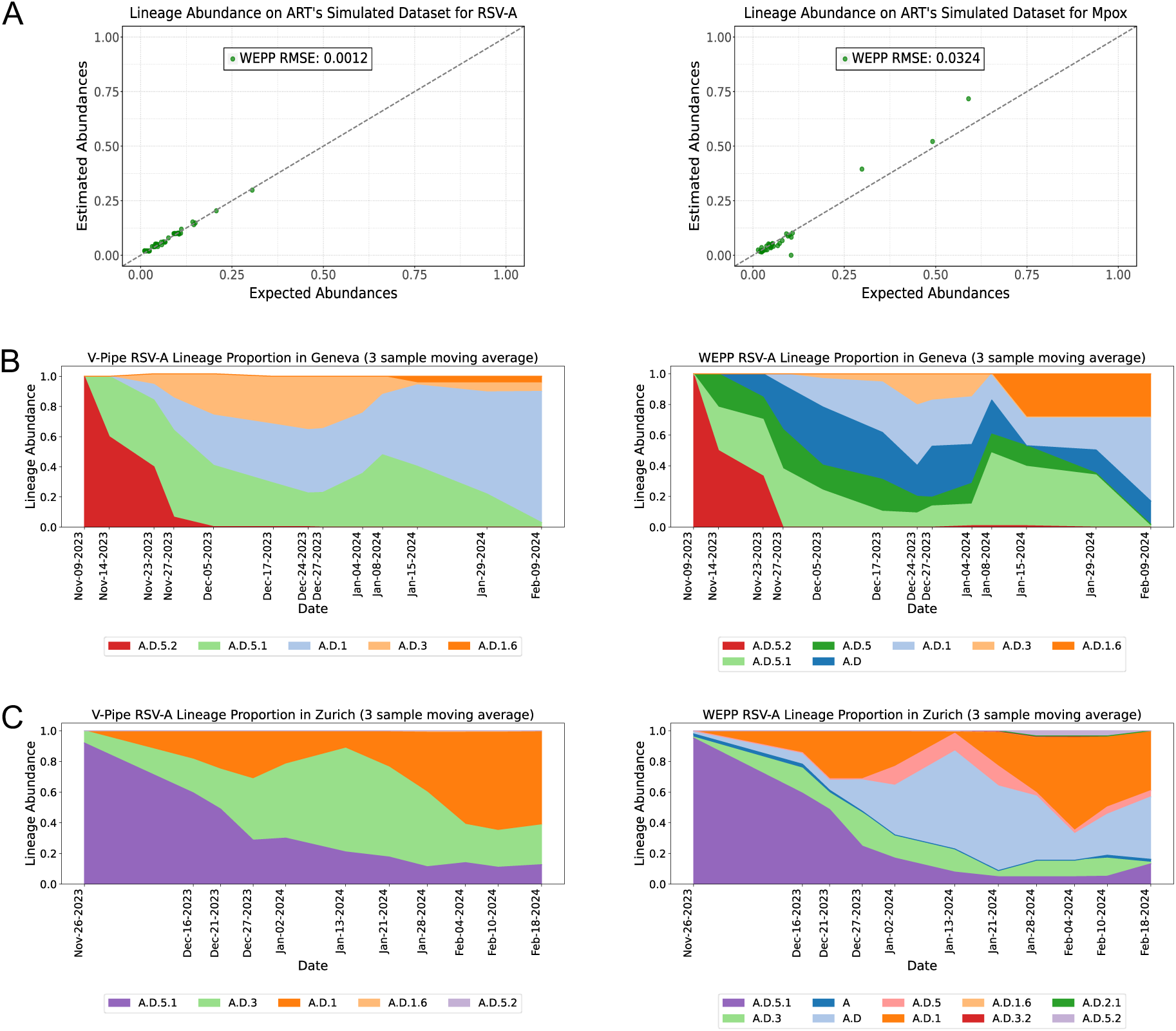
Analyzing WEPP’s lineage abundance results on non-SARS-CoV-2 pathogens. (A) Left: Panel shows WEPP’s lineage abundance accuracy on simulated RSV-A datasets. Right: WEPP’s lineage abundance results on simulated Mpox datasets. (B) Left: V-Pipe’s lineage abundance results on wastewater samples from Geneva, Right: WEPP’s abundance estimates on Geneva’s wastewater samples. (C) Left: V-Pipe’s lineage abundance results on wastewater samples from Zurich, Right: WEPP’s abundance estimates on Zurich’s wastewater samples.

We further evaluated WEPP’s performance on real RSV-A wastewater samples using data from Korne-Elenbaas et al.^34^, collected in Geneva and Zurich during the 2023–2024 RSV-A outbreak. Relative to V-Pipe^20^, the method used in the original study, WEPP consistently detected additional lineages—A.D and A.D.5—at notable proportions across multiple days in both Geneva and Zurich (Figure 5B and 5C, Supplementary Table 8). WEPP’s enhanced detection is attributed to its comprehensiveness, as it used the entire RSV-A phylogeny, whereas the V-Pipe analysis was limited to eight lineages derived from the 51 RSV-A genomes reported in Switzerland during that period. To further validate these findings, we independently analyzed a subset of samples using Freyja, which also detected the two additional lineages identified by WEPP in both locations, strengthening confidence in WEPP’s results.

These results underscore WEPP’s high sensitivity and adaptability in detecting circulating lineages of diverse pathogens from wastewater, including those missed by clinical surveillance, by leveraging both observed and inferred ancestral sequences across the entire phylogenetic tree. This approach is particularly valuable for pathogens with sparse clinical sequencing, where existing WBE tools would struggle.

## Discussion

WBE offers a powerful tool to fight local epidemics and outbreaks, and is helping mitigate the huge disparities that exist globally in clinical surveillance capabilities^60^. Although global clinical sequencing and data-sharing efforts for infectious pathogens have greatly accelerated in recent years, particularly since the onset of the COVID-19 pandemic—this massive wealth of clinical sequencing data has largely been underutilized in WBE. For example, despite over 16 million SARS-CoV-2 isolates sequenced globally to date^61^, most COVID-19 WBE studies heavily subsample this data, discarding over 99–99.9% of available clinical isolates through aggressive pre-filtering strategies^35^ or limiting searches to the roots of just a few thousand named lineages^2,21–24,26^.

Our tool, WEPP, overcomes these limitations by introducing a novel approach: it phylogenetically places wastewater sequencing reads onto a comprehensive mutation-annotated phylogeny, whose nodes represent both all global clinical sequences and their inferred ancestors. This enables WEPP to select a subset of nodes likely present in the wastewater sample. To meet the computational demands of analyzing millions of reads against tens of millions of phylogenetic nodes, WEPP integrates a suite of algorithmic and software optimizations (Methods), allowing it to perform this analysis efficiently and at scale.

In doing so, WEPP enhances WBE in three significant ways:

1. **Improved accuracy:** As demonstrated across a wide range of simulated, synthetic, and real wastewater samples, WEPP markedly improves lineage abundance estimates compared to tools that limit searches to lineage roots^2^ or apply aggressive prefiltering to select haplotypes^35^.
2. **Enhanced resolution:** By effectively leveraging mutation-annotated trees (MATs)— UShER-based phylogenies that are updated daily, annotated with lineage labels, and include both global clinical sequences and their inferred ancestors—WEPP can sensitively and precisely identify likely phylogenetic haplotypes present in wastewater. For densely sampled pathogens like SARS-CoV-2, this enables near-haplotype-level resolution, unlocking new applications such as tracking intra-lineage clusters, identifying the geographic origins of newly introduced clusters, and detecting emerging variants.
3. **Greater timeliness:** Because WEPP selects haplotypes directly from frequently updated MATs, it can bypass delays associated with the formal designation of new lineages, which often lag sequence collection by days to months^62^. This allows WEPP to rapidly detect whether newly identified clinical variant clusters from elsewhere in the world are already present in local wastewater samples. Moreover, WEPP’s reporting of unaccounted alleles enables the timely identification and analysis of cryptic or novel variants.

WEPP’s analysis can also complement several existing WBE tools. For example, WEPP can be integrated into multi-pathogen wastewater sequencing pipelines, such as the one developed by Tisza et al.^63^, to enhance the resolution of metagenomic surveillance and enable early detection of emerging variants. The unaccounted alleles identified by WEPP can also be fed into the clustering-based method developed by Amman et al.^26^ and estimate a novel variant’s reproduction numbers from multiple wastewater samples. Likewise, while WEPP reports uncertainty in haplotype detection based on the informative sites available in wastewater, its abundance estimates can be combined with methods like Dreifuss et al.^64^ to assess uncertainty in abundance quantification. Additionally, WEPP’s dashboard provides fine-grained visualizations of read mappings, unaccounted alleles, and haplotype selections within the context of global clinical data, complementing other wastewater dashboards^65–68^ that focus on broader lineage-level or viral concentration summaries. Our future efforts would involve integrating WEPP with these complementary tools. We also plan to incorporate both insertions and deletions into WEPP’s analysis with the help of recently developed PanMATs^69^, although our preliminary analysis suggests that incorporation of deletions does not improve the accuracy of lineage abundances and haplotype detection (Supplementary Results 1). We will also explore further algorithmic and acceleration techniques, such as phylogenetic placement heuristics^70^ and GPU acceleration^71,72^, to further enhance the scale and efficiency of WEPP.

As a summary, WEPP’s integration of clinically derived global phylogenetic data with wastewater surveillance enables high-resolution analysis and marks a transformative advance in pathogen monitoring. Importantly, WEPP maintains robust performance across multiple pathogens as well as on error-prone long-read data (e.g., Oxford Nanopore Technologies), achieving a 3.2-fold lower RMSE than Freyja (Supplementary Results 2). This positions WEPP to significantly strengthen and expand the scope of current wastewater surveillance efforts, enabling faster and more precise analysis of emerging threats posed by various pathogens.

## Methods

### Datasets

#### Mutation-Annotated Trees (MATs) and Pangenome Mutation-Annotated Trees (PanMATs)

The MAT used for the analysis of simulated wastewater datasets was dated December 25, 2023, and comprised publicly available sequences aggregated from GenBank, COG-UK, and the China National Center for Bioinformation (https://hgdownload.gi.ucsc.edu/goldenPath/wuhCor1/UShER_SARS-CoV-2). The tree was rooted to the GenBank reference sequence MN908947.3 and consisted of 7,840,184 sequences. In contrast, analyses of the synthetic wastewater datasets, as well as samples from Annavajhala et al.^10^ and Suarez et al.^36^, were performed using the MAT dated December 15, 2023, which additionally incorporated restricted sequences from the Global Initiative on Sharing All Influenza Data (GISAID), resulting in a total of 15,935,784 sequences. Analyses of RSV-A and Mpox datasets were conducted using publicly available sequences using the corresponding MATs dated April 25, 2025 (https://genome-test.gi.ucsc.edu/cgi-bin/hgPhyloPlace). All MATs were pre-annotated with PANGO^73^ lineage annotations.

PanMAT is a mutation-annotated tree that also stores insertions and deletions along with substitutions and uncertain alleles^69^. We constructed a PanMAT incorporating substitutions, deletions, and uncertain alleles using sequences from the MAT dated December 25, 2023, after excluding 45,144 haplotypes lacking corresponding FASTA files, resulting in a total of 7,840,184 sequences. Each sequence was pairwise aligned to the reference genome (GenBank: MN908947.3) using the Nextclade aligner^74^ that excluded insertions. The resulting MSA and MAT-derived newick tree was used to generate the PanMAT with the command: “*./panmanUtils --input-msa <file.fa> --input-newick <file.nwk> -o <file.panman> --low-mem-mode --reference Wuhan-Hu-1*”. The PanMAT was used to evaluate the impact of incorporating deletions into WEPP’s analysis (Supplementary Results 1).

### Wastewater Datasets

To comprehensively evaluate WEPP’s performance, we used a combination of simulated datasets and synthetic control mixtures with known ground truths, along with real wastewater samples. Below, we detail the construction and characteristics of each dataset.

- **Simulated wastewater datasets:** We generated in silico SARS-CoV-2 wastewater datasets using the SWAMPy simulator^46^, guided by lineage proportions observed at Point Loma between December 2022 and December 2023. Each simulated sample contained 100 haplotypes drawn proportionally from the observed lineage distributions. SWAMPy was used with the following parameters to generate 1.6 million reads: “*--primer_set n2 --n_reads 800000 --read_length 149*”, while “*--minQ 25*” was set in the “art_runner.py” script to reflect typical wastewater read quality profiles. For the samples used to mimic realistic cluster emergence scenarios, we first removed the haplotypes found (via ClusterTracker^30^) in the new region from the MAT and simulated 5% of wastewater reads from one of the removed haplotypes, while the remaining 95% were drawn from lineages circulating in the region at that time. For the experiments involving the analysis of unaccounted alleles, we used ART simulator^75^ instead of SWAMPy to ensure that the novel mutations were not dropped by the simulator. For these experiments, we created four base mixtures and simulated each with a novel haplotype at 5%, 10%, or 20% abundance, resulting in 12 samples with 1.6 million reads each. These were evaluated on phylogenetic trees where the novel haplotype and its surrounding nodes within 3, 5, or 7 mutation distances were removed, producing a total of 36 distinct test cases. As SWAMPy only supports SARS-CoV-2, ART was also employed to simulate wastewater datasets for RSV-A and Mpox. For these pathogens, we generated samples containing 10, 20, or 50 randomly selected haplotypes.
- **Synthetic control datasets**: To compare WEPP and Freyja’s performance on synthetic controlled mixtures, assay-ready RNA controls from Twist Biosciences (https://www.twistbioscience.com/products/ngs/synthetic-viral-controls?tab=sars-cov-2-controls) were spiked into SARS–CoV–2–negative wastewater to generate synthetic samples. We benchmarked WEPP on both Illumina-sequenced mixtures prepared by the Indiana Department of Health (IDOH) and Oxford Nanopore Technologies (ONT)-sequenced mixtures from Ferdous et al. ^45^, to assess generalizability across sequencing technologies. The IDOH samples were produced by creating five different synthetic mixtures; three of which contained three haplotypes in equal proportions, while the remaining two contained a single haplotype. Each sample was independently prepared and sequenced twice to assess wet-lab reproducibility, yielding 10 samples in total. Additionally, WEPP was tested on three ONT-sequenced synthetic mixtures from Ferdous et al., each comprising eight distinct haplotypes in varying proportions. Individual haplotype abundances ranged from as low as 6.25% to as high as 31.25%, allowing us to assess WEPP’s performance under challenging abundance distributions and sequencing noise.
- **Real wastewater samples:** To evaluate WEPP’s performance on real-world data, we analyzed 77 wastewater samples from four different sources to assess a range of surveillance goals: correlation with clinical sequencing, early detection of novel SARS-CoV-2 variants and mutations, cryptic lineage detection, and generalizability to non-SARS-CoV-2 pathogens.

1. **SARS-CoV-2 wastewater from Point Loma, San Diego (22 samples)** We analyzed 22 samples collected between November 27, 2021, and February 7, 2022, from San Diego’s largest wastewater treatment plant, as described in Karthikeyan et al.^2^ (BioProject Accession ID: PRJNA819090). To mimic real-time surveillance, each sample was analyzed using MATs that also included private sequences from GISAID, which were constructed before the sample’s collection date. Specifically, samples collected before December 1, 2021, used the November 15, 2021, MAT; subsequent samples used MATs that were updated every 15 days. WEPP’s lineage abundance estimates were compared with those from Freyja and “oracle” clinical sequencing data. Unaccounted alleles identified by WEPP were also compared against all sequences reported from San Diego from November 2021 to March 2022, to assess their rarity and potential cryptic mutations. We also analyzed the WEPP inferred haplotypes and compared them against all clinical sequences from San Diego collected during the same period, along with their earliest detection dates, to evaluate how accurately and early WEPP could detect these sequences from wastewater.
2. **SARS-CoV-2 samples from NewYork-Presbyterian Hospital, Columbia University Irving Medical Center (13 samples)** We analyzed 13 wastewater samples collected in December 2022 from the NewYork–Presbyterian Hospital at Columbia University Irving Medical Center, from Annavajhala et al.^10^ (BioProject Accession ID: PRJNA1230707). WEPP’s and Freyja’s lineage abundance estimates were compared against clinical sequences obtained from the same hospital and time.
3. **Cryptic lineage containing SARS-CoV-2 samples from Suarez et al. (18 samples)** We evaluated 18 wastewater samples containing different cryptic SARS-CoV-2 lineages as described in Suarez et al.^36^. For each sample, we focused on “cryptic lineage– defining amino acid substitutions” as reported in the original study. These were translated into nucleotide-level mutations from their data, and in case of multiple codons encoding the same amino acid change, we selected the nucleotide mutation least frequent in WEPP’s inferred haplotypes. We then compared these mutations against WEPP’s unaccounted allele calls and also analyzed them across different WEPP’s thresholds, namely coverage depth and deviation in depth-weighted allele frequency from expectation.
4. **RSV-A samples from Geneva and Zurich (24 samples)**

To test WEPP’s generalizability, we analyzed 24 RSV-A samples collected during the 2023–2024 outbreak from wastewater in Geneva and Zurich, as shared by De Korne-Elenbaas et al.^34^ (ENA Study Accession: PRJEB85787). WEPP’s lineage abundance estimates were then compared to those reported by V-Pipe, which was restricted to lineages previously observed in Swiss clinical sequences during the outbreak period^34^.

### Baseline Tools

We compared WEPP’s performance against two baselines: Freyja^2^ and the EM algorithm proposed by Pipes et al.^35^. Freyja has consistently been ranked among the most accurate WBE tools in multiple benchmarking studies^45,76^ and is also widely used in practice^10,13,49,77,78^. The EM algorithm by Pipes et al. is the only other method that estimates haplotype abundances from wastewater sequencing data.

We integrated Freyja into WEPP and, when benchmarking against it, used lineage roots as input in place of phylogenetically selected haplotypes (see our implementation at https://github.com/TurakhiaLab/WEPP/tree/wepp_freyja). For the EM baseline, we refactored and reimplemented the algorithm from Pipes et al. in C++ to improve computational efficiency (see our implementation at https://github.com/TurakhiaLab/WEPP/tree/wepp_em), applied a default haplotype filtering threshold of 1% allele frequency and sequencing error rate of 0.5%, as described in their paper. Inputs to the EM algorithm were generated by computing mutation differences between sequencing reads and MAT sequences. The algorithm was terminated when the log-likelihood change between successive iterations dropped below 0.1% or after a maximum of 25 iterations.

### WEPP: Pipeline Overview

The WEPP pipeline is a Snakemake workflow that consists of three main components: i) Quality Control filtering and Read Subsampling, ii) WEPP Algorithm, and iii) WEPP Dashboard analysis. The pipeline requires as input: i) raw wastewater sequencing reads, ii) mutation-annotated tree, iii) corresponding reference genome, and iv) BED file specifying primer locations.

### Quality Control Filtering and Read Subsampling

The quality control begins by aligning raw sequencing reads to the reference genome using minimap2^79^. Aligned reads are then processed with iVar^80^ for primer trimming based on the provided BED file. WEPP then applies two layers of allele-level quality control to ensure reliability in downstream analysis. First, bases with a Phred quality score below a user-defined threshold (MIN_Q, default: 20) are masked within the reads. Second, alleles with observed frequencies below a specified minimum allele frequency (MIN_AF, default: 0.5% for Illumina reads) are also masked in the reads. Importantly, all reads are retained in the dataset, but their sites failing these QC filters are masked and excluded from downstream analysis.

WEPP includes an optional read subsampling feature to reduce the runtime. It generates *N* independent subsets (default: 1000 subsets), each containing a user-specified number of randomly selected reads and selects the subset whose allele frequency distribution has the lowest Kullback-Leibler (KL) divergence from the full dataset, ensuring minimal distortion of information.

### WEPP: Algorithm Details

WEPP consists of three main processing stages: (i) Read Mapping and Haplotype Selection, (ii) Neighbor Addition, and (iii) Abundance Estimation (Figure 1C). Once WEPP infers the haplotypes, unaccounted alleles are computed, and reads are mapped to these inferred haplotypes. The core algorithm is implemented as multi-threaded C++ using the oneTBB library^81^, and is integrated with the Quality Control pipeline and interactive WEPP Dashboard via a Snakemake workflow. Detailed implementation of different stages is described below.

1. **Read Mapping and Haplotype Selection:** WEPP processes all sequencing reads after the quality control stage and identifies genome sites that remain uncovered. It performs haplotype grouping by clustering haplotypes that differ only at these uncovered positions and reports them as ‘Uncertain Haplotypes’ for each selected representative haplotype (Figure 1B(i)). For each read, WEPP computes two values: the parsimony^82^ (i.e., minimum number of mutations required to place a read on the phylogenetic tree) and the number of equally parsimonious placements (EPPs). Both of these values are then used to assign a weight to every read. Once all reads are mapped to the tree, WEPP assigns a score to each node by summing the weights of all reads that map parsimoniously to it. This score is then regularized by the fraction of the genome covered by those reads. Next, WEPP applies a greedy haplotype selection algorithm. It begins by selecting the node with the highest score as the initial haplotype. After the haplotype is selected, the scores of all remaining nodes are updated by subtracting the weights of reads that map parsimoniously to the selected haplotype. This process of haplotype selection followed by score update is repeated iteratively until all reads are either exhausted or a predefined number of haplotypes are selected (default: 300).

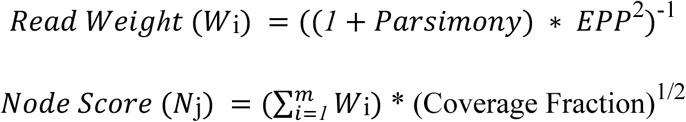

**2. Neighbor Addition:** After identifying an initial set of haplotypes using the greedy haplotype selection algorithm, WEPP expands this candidate set by including neighboring nodes around each haplotype. The mutation radius used to define these neighbors is chosen adaptively to maintain the total size of the final candidate set at a fixed level (default: 5,000 haplotypes). To further constrain the search space, no more than a fixed number of neighbors (default: 50) are added per initially selected haplotypes. Neighbor addition ensures that closely related haplotypes, which differ by only a few mutations, are not overlooked.
**3. Abundance Estimation:** WEPP applies an extended version of Freyja’s abundance estimation framework to the expanded haplotype set generated through Neighbor Addition. To reduce overfitting and noise from spurious matches, haplotypes with low abundance estimates (default: 0.5%) are greedily pruned. Starting with the lowest-abundance haplotype, each haplotype below the threshold is removed and its proportion reassigned to the closest neighbor from the candidate set. After all remaining haplotypes are above the threshold, Freyja is rerun to refine the estimates, and any newly sub-threshold haplotypes are discarded. This greedy approach avoids the pitfall of naively applying the abundance threshold, where an entire haplotype cluster gets eliminated if all its members fall below the threshold, despite collectively representing a significant abundance. By reassigning proportions before removal, WEPP preserves the cluster’s representation and improves robustness. As illustrated in Figure 1C, WEPP then enters an iterative refinement loop, where neighbors of the retained haplotypes are re-added to the candidate pool (default: mutation distance radius of 2; maximum 500 neighbors per haplotype), and the abundance estimation step is repeated. This process continues until the retained haplotype set converges (i.e., remains unchanged over two successive iterations) or a maximum number of iterations (default: 10) is reached.
**4. Unaccounted Alleles:** WEPP extends Freyja’s optimization framework to identify *Unaccounted Alleles*, alleles present in wastewater but not explained by the selected haplotypes. While Freyja minimizes the depth-weighted absolute differences between observed and predicted allele frequencies (*weighted freq_diff*), WEPP builds on it by first computing the mean and standard deviation of these differences across all mutations. Mutations that significantly deviate from this distribution are flagged as unaccounted, with their direction of deviation indicating whether the allele is over- or under-represented in the selected haplotypes. A mutation is labeled as an unaccounted allele if it satisfies all of the following criteria:
  a. *weighted freg_diff_i_* ∉ [*mean* − 2 ∗ *std deviation, mean* + 2 ∗ *std deviation*]
  b. *Depth_i_* > 0.6 ∗ *mean Depth*
  c. |*freq_diff_i_*| /*Observed_freq_i_* > 0.5
5. **Read-to-Haplotype Mapping and Potential Haplotypes for unaccounted alleles:** After identifying all haplotypes present in a sample, WEPP performs parsimony-based mapping of each read to this haplotype set to determine its most likely origin. To associate each unaccounted allele with a potential haplotype, WEPP first masks the unaccounted alleles in the reads and groups the reads based on the specific unaccounted allele they contain. Within each group, it identifies the haplotype that receives the highest number of parsimonious mappings and records it as the most likely source of that unaccounted allele.

### WEPP: Optimization Details

Mapping more than 4 million sequencing reads from wastewater to a phylogenetic tree of ∼16 million SARS-CoV-2 sequences is computationally challenging. To make this process feasible in near real-time, WEPP employs several key optimizations:

1. **De-duplication of reads.** WEPP first identifies and collapses duplicate reads, performing parsimony-based mapping only for this unique set. Node scores are then adjusted by the frequency of each unique read in the dataset, greatly reducing redundant computations.
2. **Parallelized read mapping:** WEPP extracts parallelism during the read mapping stage by assigning each read to a separate thread, allowing multiple reads to be processed simultaneously. This thread-level parallelism significantly accelerates the search and improves scalability with a large number of unique reads.
3. **Phylogenetic tree condensation.** To minimize unnecessary traversal and track ‘Uncertain Haplotypes’ (Figure 1B(i)), WEPP prunes the global phylogenetic tree by retaining only those nodes with mutations at positions covered by the wastewater reads. It stores the condensed node information at each retained node, resulting in a much smaller and more relevant subtree that significantly accelerates the mapping process.
4. **Region-aware tree search.** WEPP partitions the reads based on the genome segment they cover and constructs localized subtrees only containing mutations specific to those regions. This region-aware approach avoids checking unrelated nodes with no relevant mutations, leading to faster and more efficient parsimony computation.

### Evaluation metrics

We evaluated the performance of WEPP against baseline methods using simulated and synthetic control mixtures. Comparisons were based on three key metrics: Lineage Abundance RMSE, Weighted Haplotype Distance, and Weighted Peak Distance.

- **Lineage Abundance RMSE**: Measures the root mean squared error between the expected and estimated lineage abundances.
- **Weighted Haplotype Distance**: Assesses how well the most abundant haplotypes in the sample are recovered by each tool, with greater weight given to haplotypes of higher proportion. Each true haplotype is matched to the closest estimated haplotype based on minimum mutation differences. This metric reflects *sensitivity*, lower values indicate better recovery of true haplotypes.
- **Weighted Peak Distance**: Captures *precision* by evaluating how closely the highest estimated abundance haplotypes reported by a tool match true haplotypes present in the wastewater sample. Lower values of this metric indicate fewer spurious inferred haplotypes.

The weighted distance metrics can be defined more formally by assuming H={h_1_,h_2_,…,h_n_} is the set of true haplotypes in the sample with corresponding proportions wh, and P={p_1_,p_2_,…,p_m_} is the set of inferred haplotypes with corresponding proportions wp. The mutation distance captures the number of single-nucleotide substitutions separating the true haplotype h_i_ from the inferred haplotype p_j_, which is denoted by Dist(h_i_,p_j_).

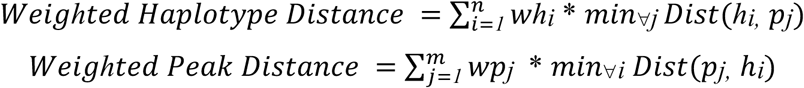

### Execution Environment and Runtimes

All experiments were performed on an Intel Xeon Silver 4216 processor with 768GB DDR4 memory and 4.0TB SATA. WEPP was run using 48 CPU threads, with runtimes varying from a few minutes to 14 hours depending on the size of the MAT and the number of reads in the sample. For example, the RSV-A datasets from Korne-Elenbaas et al.^34^ contained approximately 2 million sequencing reads, which were analyzed in under 10 minutes, since the RSV-A MAT contained only 8,215 sequences. In contrast, Point Loma wastewater samples were analyzed in approximately 4 hours, containing a similar number of reads due to a much larger SARS-CoV-2 MAT comprising 7.45 million publicly available sequences. The longest runtimes were observed for synthetic samples from IDOH, which were analyzed using an even larger MAT that also included restricted GISAID sequences, increasing its size by 2.14-fold (15.93 million sequences). Additionally, these IDOH samples contained about 25% more unique reads than the Point Loma samples, even after subsampling to 1 million reads, which limited the effectiveness of certain WEPP optimizations (see above).

### WEPP Dashboard: Overview and Implementation Details

To enhance the utility and interpretability of WEPP results, we developed an interactive visualization dashboard using the Taxonium framework^44^ and a custom JBrowse plugin^83^, which enables researchers and public health officials to interpret wastewater surveillance results more effectively. The dashboard displays detected haplotypes, their corresponding lineages, along with associated abundances and mutation profiles in an intuitive visual format. This would enable public health officials to rapidly identify emerging haplotype clusters and lineages, monitor shifts in their prevalence over time, and make informed, data-driven decisions for targeted community health interventions (Figure 1B).

The dashboard customizes the Taxonium framework^44^ for visualizing detected haplotypes within their evolutionary context. This interactive component enables users to navigate the complete phylogenetic tree through intuitive pan and zoom controls while accessing contextual information by hovering over specific nodes. The dashboard’s search panel displays detected haplotypes with their corresponding abundances, lineage classifications, and associated uncertain haplotypes, enabling rapid identification of variants of interest (Figure 1B(i)).

When users select an annotated node on the dashboard, a custom JBrowse plugin^83^ is triggered to enable high-resolution sequence analysis. This plugin provides an interactive, multi-track genome browser view that includes: (i) annotated gene features along the reference genome, (ii) reconstructed haplotype sequences aligned to the reference, and (iii) individual sequencing reads from the wastewater sample, mapped to both the reference and haplotype sequences. This layered visualization enables users to explore how specific mutations are supported by sequencing reads and to assess their linkage within the context of gene structure and inferred haplotypes (Figure 1B(ii)). Additionally, users can click on any individual sequencing read within the BAM track to explore alternative haplotype associations and view unaccounted alleles (Figure 1B(iii)). Similarly, clicking on a haplotype sequence reveals a list of all potential unaccounted alleles linked to that haplotype, along with the associated uncertain haplotypes (Figure 1B(iv)), which aid in the identification of low-frequency or ambiguous variant signals.

To visualize results on this dashboard, the WEPP’s pipeline generates two annotated BAM files. The first contains WEPP-inferred haplotypes, with metadata added to the BAM header using read groups (@RG) to identify each unique haplotype. These read groups include tags such as HS (haplotype proportion), HL (lineage label), and UH (uncertain haplotype flag). The second BAM file contains the original wastewater sample reads, annotated with tags like UM to flag unaccounted alleles, RG to link a read to its possible source haplotypes, and EP to indicate the number of possible haplotype sources. Both BAM files also include @CO comment lines, each of which describes an uncertain allele with a unique identifier, its position and base, associated residue, frequency, and depth.

The codebase for this dashboard is hosted on a dedicated public repository (https://github.com/pratikkatte/Wastewater-Dasboard), which is included as a submodule within the main WEPP repository (https://github.com/TurakhiaLab/WEPP).

## Data Availability

All data produced in the present study are available upon reasonable request to the authors.

## Acknowledgments

We gratefully acknowledge all data contributors—namely, the authors and their originating laboratories responsible for specimen collection, and the submitting laboratories for generating and sharing sequence data and metadata via GISAID, GenBank, COG-UK, and the China National Center for Bioinformation—on which this research is based. We thank David Schaeper and the Indian Department of Health for providing the synthetic wastewater datasets. We are also grateful to the authors of Ferdous et al.^45^, particularly Samuel Kunkleman, for assistance in accessing their benchmarking data. We thank Medini Annavajhala for sharing both wastewater and clinical samples from her study. We are deeply appreciative of Angie Hinrichs for providing access to private MATs containing GISAID sequences, along with her technical guidance and insightful feedback. We also thank Kristian Andersen, Joshua Levy, Karthik Gangavarapu, Russell Corbett-Detig, Jason Caravas, and Daniel Cornforth for their valuable feedback. We acknowledge Sumit Walia for his contributions in generating the PanMAT and helping it integrate into WEPP. This work was supported by funding from the U.S. Centers for Disease Control and Prevention through the Office of Advanced Molecular Detection (CDC contract #75D30123C17463), the Amazon Research Award (Fall 2022 CFP), and funding from the Hellman Fellowship.

## Supplementary Material

### Supplementary Results 1: Incorporating deletions via PanMAT in WEPP has a negligible impact on SARS-CoV-2 variant detection accuracy

Some recently proposed WBE tools, such as Lineagespot^24^ and QuaID^5^, incorporate deletions in addition to substitutions to detect variants from wastewater. The authors of QuaID claimed that the reduced sensitivity of Freyja, which only considers substitutions in detecting emerging variants of concern, could be due to its lack of consideration of deletions. However, to our knowledge, no study has systematically quantified the impact of deletion inclusion alongside substitutions in SARS-CoV-2 variant detection from wastewater.

We evaluated the impact of considering deletions and substitutions in WEPP by using PanMAT^69^, a generalized form of MATs that can represent insertions, deletions, and complex mutations in addition to substitutions. To perform this analysis, we constructed a PanMAT that included deletions and substitutions from nearly eight million public SARS-CoV-2 sequences available up to December 25, 2023 (Methods). We excluded insertions for this analysis as it would have required us to define a non-reference-based coordinate system, which would make the analysis quite complex. We then compared WEPP-PanMAT (modified WEPP to support PanMATs), which includes both substitutions and deletions, against WEPP-MAT (default WEPP that uses MATs), which only considers substitutions, on SWAMPy simulated datasets and assessed their accuracy and resolution in variant detection.

The results, shown in Supplementary Figure 4B, indicate that incorporating deletions through PanMAT did not improve any of the performance metrics on the SWAMPy simulated data. We attribute this to the relative rarity of deletions compared to substitutions in SARS-CoV-2 sequences, as illustrated in Supplementary Figure 4A. On average, we find that there are nearly 14 times more substitutions than informative deletions between adjacent lineages (parent and child). These results suggest that deletions contribute minimal additional signal for variant detection, as the numerous substitutions present in the MATs already capture the key distinguishing genomic differences between lineages, and according to Walia et al.^69^, the insertions tend to be even fewer. In other words, our preliminary findings suggest that incorporating insertions and deletions into the WEPP analysis is unlikely to provide major gains.

### Supplementary Results 2: WEPP generalizes to noisy long-read sequencing data

Wastewater sequencing protocols vary widely across regions and institutions, particularly in terms of library preparation methods and sequencing platforms. This variability makes it essential for any WBE tool to be broadly generalizable and effective across different settings. To evaluate WEPP’s robustness, we tested its performance on Oxford Nanopore Technologies (ONT) long-read sequencing data, which are known to have higher error rates than short-read platforms.

We analyzed three synthetic control mixtures from Ferdous et al.^45^, each containing unique combinations of eight known haplotypes spiked into SARS-CoV-2 negative wastewater RNA backgrounds at varying proportions. As shown in Supplementary Figure 1A (Supplementary Table 1), WEPP achieved a 3.2-fold lower lineage abundance RMSE compared to Freyja, demonstrating strong performance despite the higher noise associated with ONT sequencing.

### Supplementary Figures

**Supplementary Figure 1:**
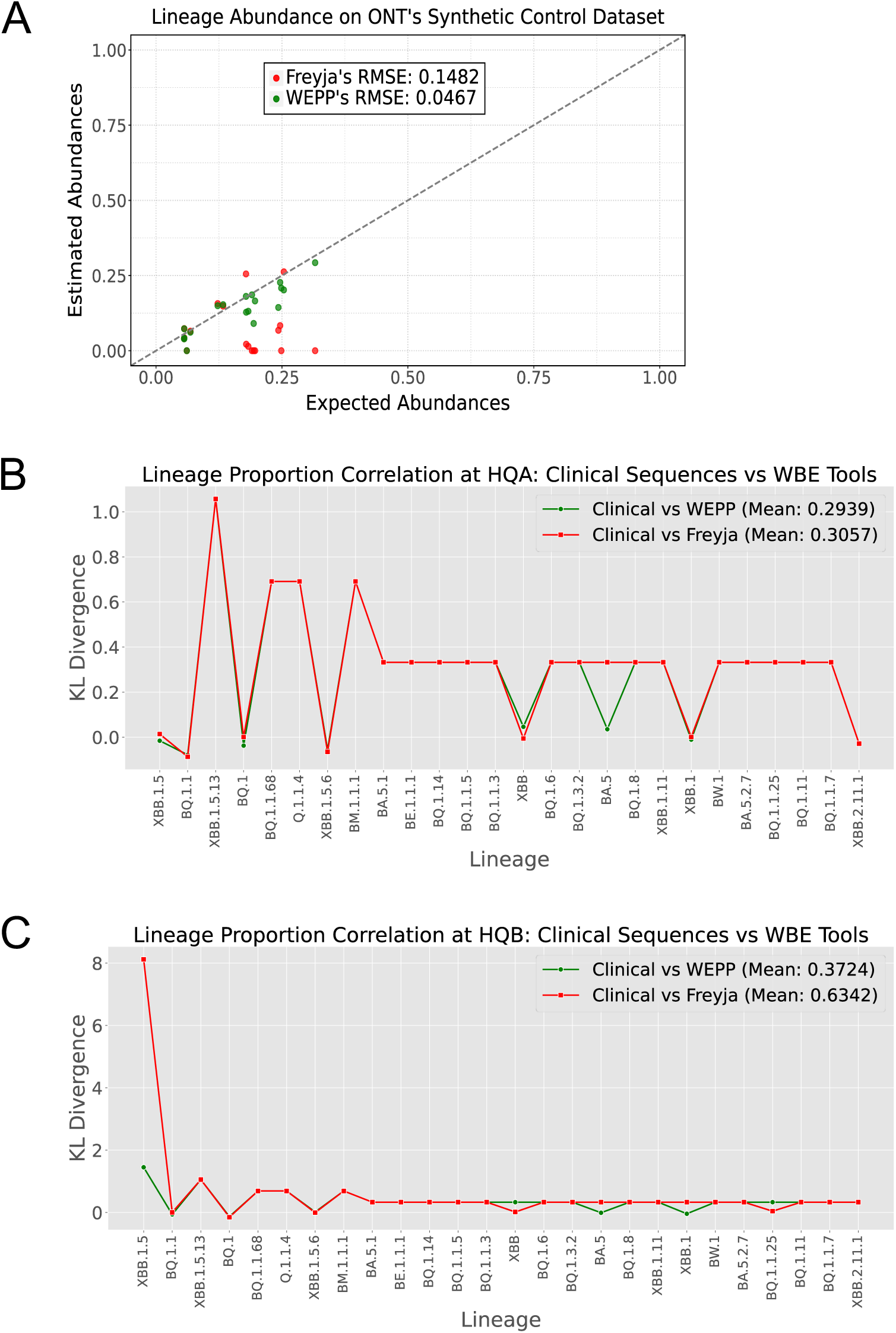
Comparison of lineage abundance between WEPP and Freyja on ONT sequenced data. (A) WEPP and Freyja’s lineage abundance comparison on the ONT sequenced synthetic control dataset. (B) Average lineage abundance estimates from wastewater samples collected in December 2022 from Hospital Quadrant A, compared with clinical sequences of hospitalized patients in the same hospital and time period^10^. (C) Average lineage abundance estimates from wastewater samples collected in December 2022 from Hospital Quadrant B, compared with clinical sequences of hospitalized patients in the same hospital and time period^10^.

**Supplementary Figure 2:**
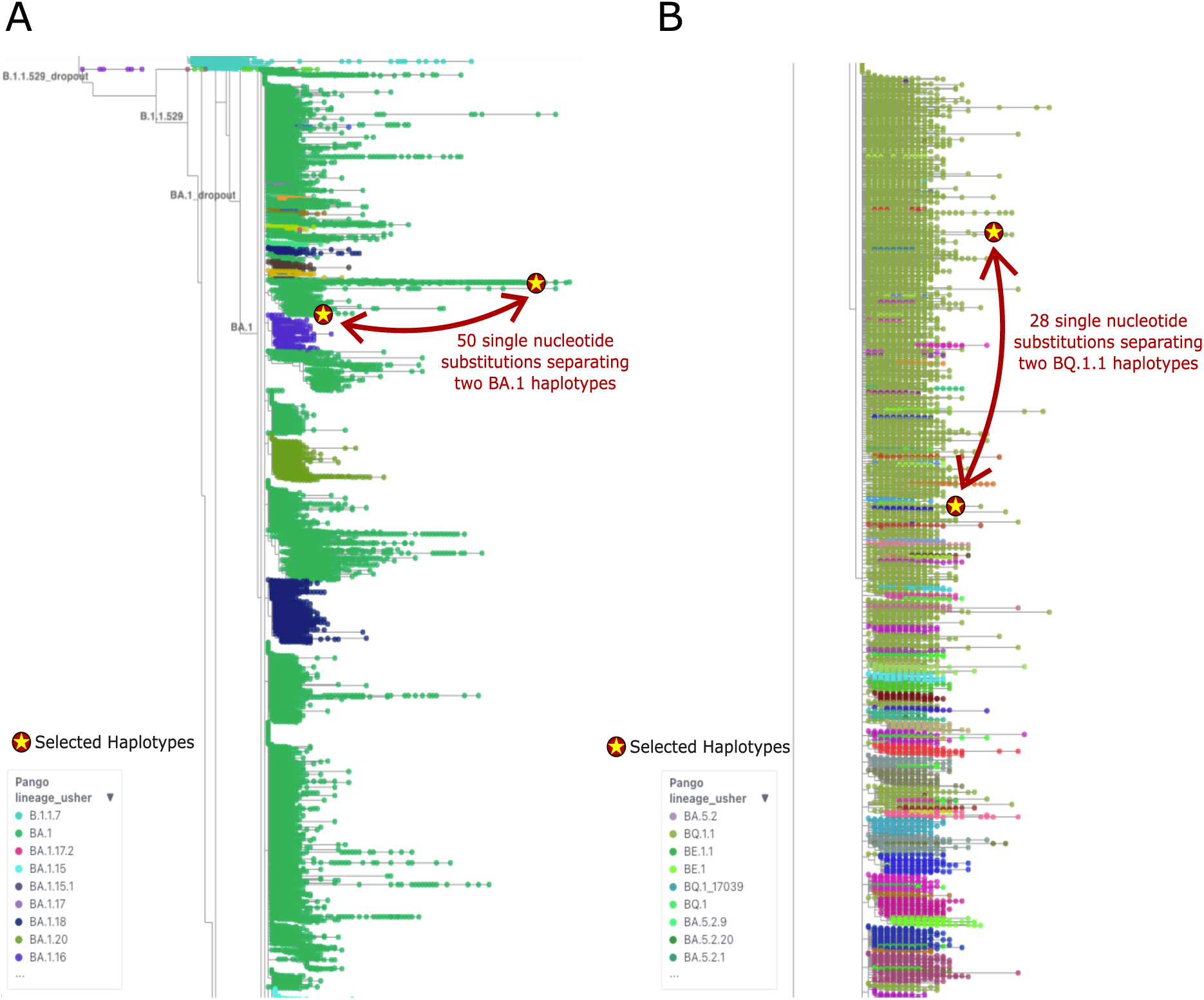
Single-nucleotide substitutions separating two haplotypes in a lineage. (A) Haplotypes “England/PHEC-5U048Z3E/2022|OX781526.1|2022-01-29” and “England/MILK-3777F8F/2022|OW112938.1|2022-02-18” belonging to lineage BA.1, (B) Haplotypes “Germany/IMS-10116-CVDP-91FBB221-5948-4641-8459-88ABFE0F8DE1/2023|OY271077.1|2023-03-06” and “USA/PA-CDC-QDX46010440/2023|OQ419762.1|2023-01-24” belonging to BQ.1.1 (right panel)

**Supplementary Figure 3:**
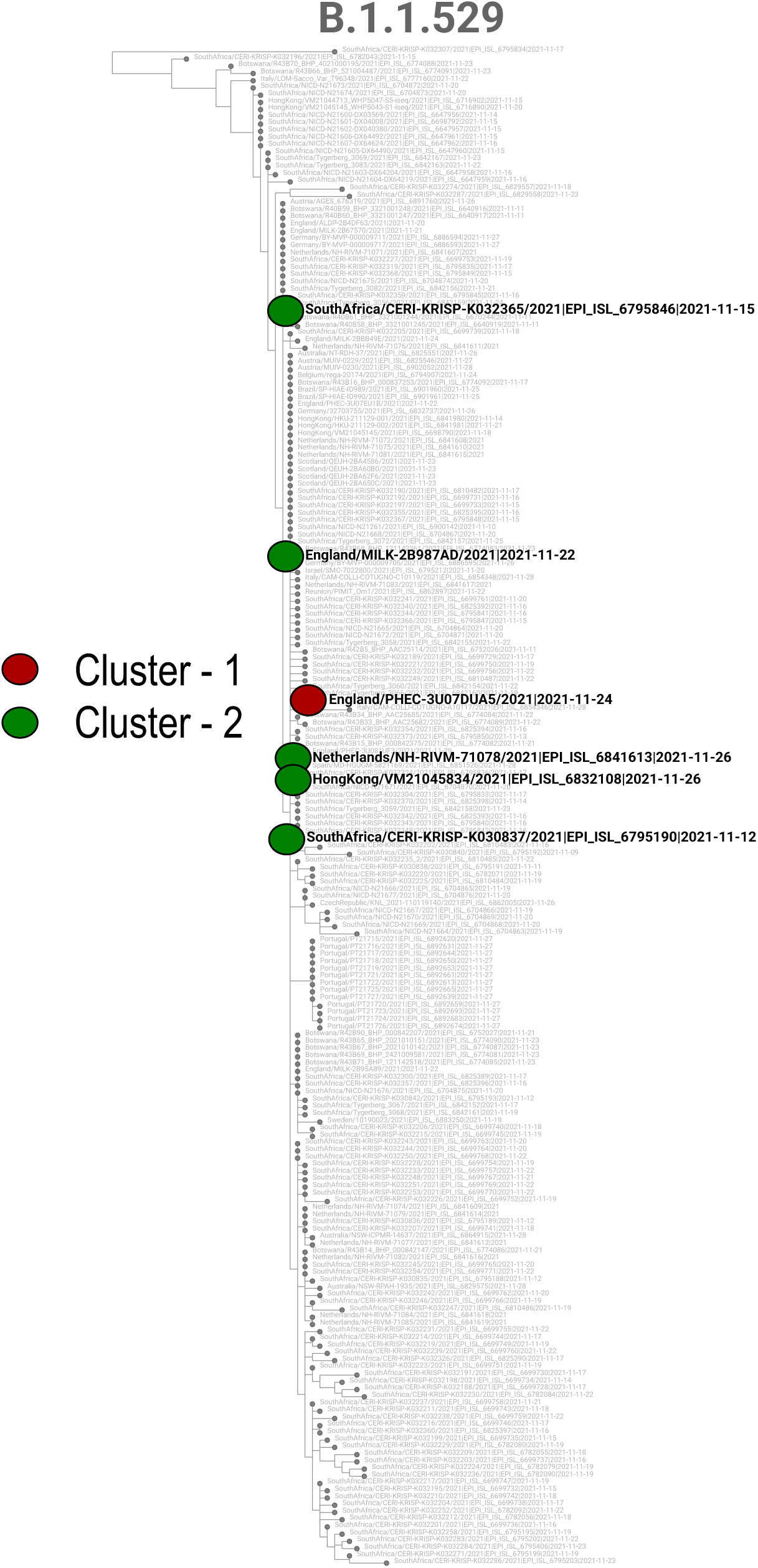
B.1.1.529 (Omicron) Haplotypes detected by WEPP from Point Loma (San Diego) wastewater samples dated December 1, 2, and 5, 2021.

**Supplementary Figure 4:**
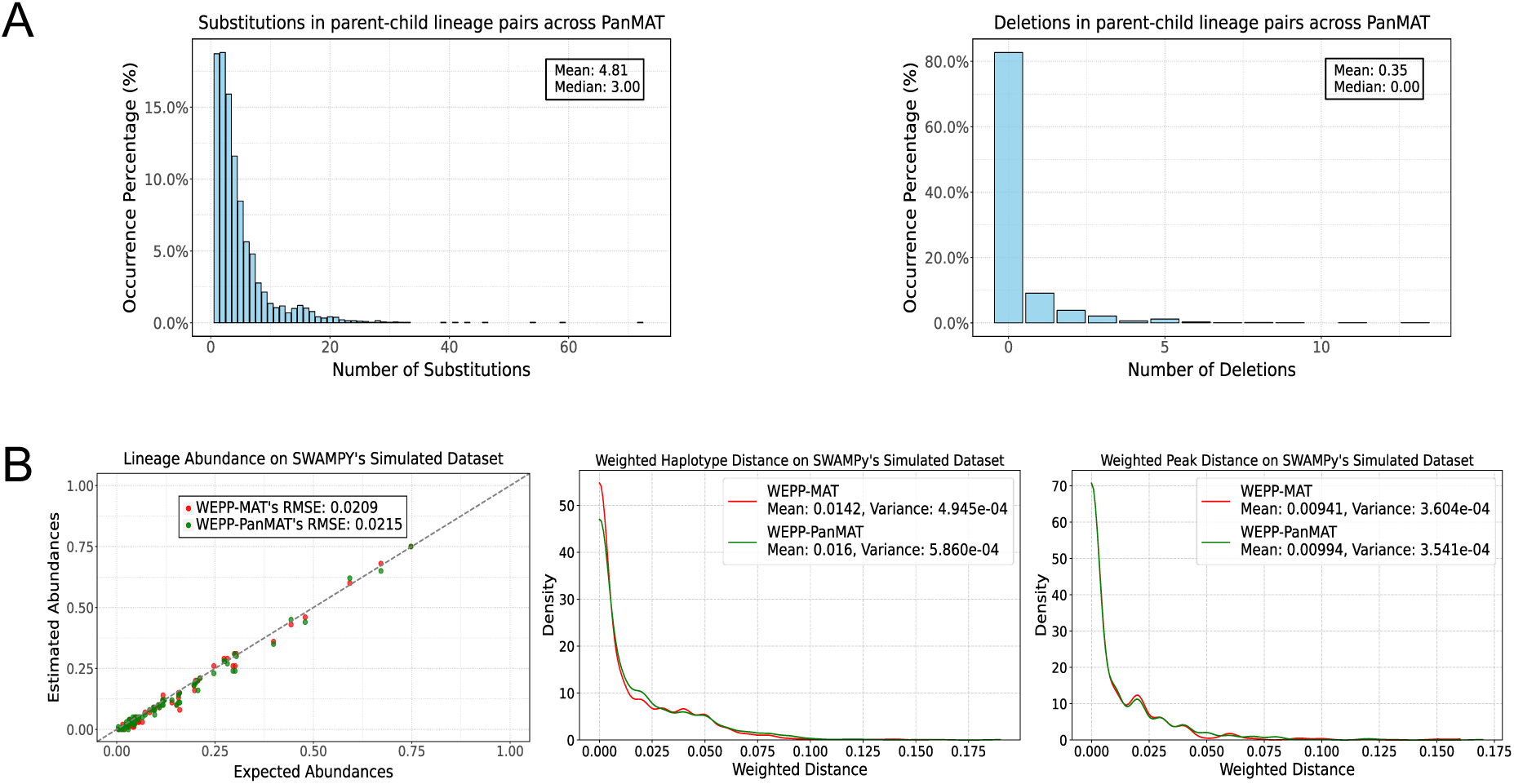
Impact of incorporating deletions in WEPP. (A) Substitution and deletion differences between parent-child lineage pairs in a PanMAT constructed with eight million public sequences available until December 25, 2023. (B) Performance comparison of WEPP using MAT (WEPP-MAT), which considers only substitutions, and WEPP using PanMAT (WEPP-PanMAT), which includes both substitutions and deletions, based on lineage abundance RMSE (left), Weighted Haplotype Distance (middle), and Weighted Peak Distance (right) on SWAMPy simulated datasets.

